# Bayesian Spatio-Temporal Modeling of COVID-19: Inequalities on Case-Fatality Risk

**DOI:** 10.1101/2020.08.18.20171074

**Authors:** Gina Polo, Carlos Mera Acosta, Diego Soler-Tovar, Julián Felipe Porras Villamil, Natalia Polanco Palencia, Marco Penagos, Juan Meza Martinez, Juan Nicolás Bobadilla, Laura Victoria Martin, Sandra Durán, Martha Rodriguez, Carlos Meza Carvajalino, Luis Carlos Villamil, Efrain Benavides Ortiz

## Abstract

The ongoing outbreak of COVID-19 challenges health systems and epidemiological responses of all countries worldwide. Although mitigation measures have been globally considered, the spatial heterogeneity of its effectiveness is evident, underscoring global health inequalities. Using Bayesian-based Markov chain Monte Carlo simulations, we evidenced that factors contributing to poverty are also risk factors for COVID-19 case-fatality, and unexpectedly, their impact on the case-fatality risk is comparable to that produced by health factors. Additionally, we confirm that both case-fatality risk and multidimensional poverty index have a heterogeneous spatial distribution, where the lastest consists of health, educational, dwelling, and employment dimensions. Spatio-temporal analysis reveals that the spatial heterogeneity in case-fatalities is associated with the percentage contribution of the health (RR 1.89 95%CI=1.43-2.48) and dwelling (RR 2.01 95%CI=1.37-2.63) dimensions to the multidimensional poverty, but also with the educational (RR 1.21 95%CI=1.03-1.49), and employment (RR 1.23 95%CI=1.02-1.47) dimensions. This spatial correlation indicates that the case-fatality risk increase by 189% and 201% in regions with a higher contribution of the health dimension (i.e., lack of health insurance and self-reporting) and dwelling dimension (i.e., lack of access to safe water, inadequate disposal of human feces, poor housing construction, and critical overcrowding), respectively. Furthermore, although a temporal decrease is evident, the relative risk of dying by COVID-19 in Colombia is still 200% higher than the established case-fatality risk based on the COVID-19 dynamics in Italy and China. These findings assist policy-makers in the spatial and temporal planning of strategies focused on mitigating the case-fatality risk in most vulnerable communities and preparing for future pandemics by progressively reducing the factors that generate health inequality.

## 1. Introduction

The ongoing outbreak of COVID-19 challenges health systems and epidemiological responses of all countries worldwide since its first report in Wuhan City, Hubei Province of China.[1] The number of human deaths associated with COVID-19 exceeds past coronavirus outbreaks,[2] and this number is increasing, mainly in the Americas. Several mitigation measures oriented to reduce the severity, named case-fatality rate (proportion of persons with COVID-19 who die from this disease), have been considered by various governments, as the household-based prevention model, which usually includes remote work and self-quarantine. Apparent differences in the COVID-19 case-fatality rate between regions[1] and temporal variations concerning the basic reproduction number *R_t_*[3] are indicators of the spatiotemporal heterogeneity of the mitigation measures and their effects. Indeed, the effectiveness of the mitigation measures depends on the time of its implementation,[4] and presumably, on other factors such as the proportion of individuals unable to working remotely, e.g., socioeconomically disadvantaged people who mostly depend on informal activities.

Besides the dependence of COVID-19 on biological, clinical, and epidemiological aspects,[5] in this work, we will show that even in populations under similar mitigation measures, COVID-19 case-fatalities can be spatially heterogeneous. This anisotropy can intuitively be associated with barriers to health services, such as hospitals and intensive care units, however, other social and economic aspects could also cause health inequalities.[6-8] Thus, we provide quantitative evidence that socioeconomic factors are as relevant as the barriers to health services for health inequalities in the COVID-19 outbreak, i.e., the systematic, avoidable and unfair differences in health outcomes that can be observed between populations, social groups within the same population or as a gradient across a population ranked by social position.[9] Based on Bayesian non-spatio-temporal and spatio-temporal modeling, we evidenced multidimensional poverty risk factors namely educational, childhood/youth, employment, health, and dwelling poverty factors, that significantly affect the inequalities on COVID-19 case-fatality risk.

Identify the factors that trigger health inequalities on the COVID-19 outbreak and its distribution over space described in terms of proximity, distance, clustering, and concentration, is fundamental to reduce the COVID-19 impact on vulnerable communities and enhance the effectiveness of mitigation measures through the implementation of individualized and effective strategies. Thus, our findings assist policy-makers in the spatial planning of strategies focused on mitigating case-fatalities in most vulnerable communities and preparing for future pandemics by progressively reduce the factors that generate health inequality.

## 2. Methods

### 2.1. Data

We use the COVID-19 death official reports by the National Institute of Health of Colombia (INS).[10] This study region has a population of around 50 million people distributed in 32 administrative units called departments and Bogotá D.C as the capital district. Colombian COVID-19 data can be interactively displayed on this dashboard (**https://rpubs.com/gppoloi/CoCo**). The first death by COVID-19 in Colombia was reported on March 16, 2020, and in total, 13837 people have died, as the INS report of August 12, 2020. Data were smoothed considering a weekly calibration and aggregated at the departmental level, and all the departments with deaths reported from week 12 (from March 16) to week 31 (to August 02) of 2020 were considered. For this reason, six eastern departments were unconsidered. Thus, data summarising population-level deaths were spatiotemporally aggregated to a set of 27 non-overlapping areal units for 20 consecutive time intervals, in this way comprising a *K* × *N* matrix of spatio-temporal observed deaths.

As proxy covariates of case fatality inequalities, we consider the Colombian Multidimensional Poverty Index (MPI) database.[9] The MPI is a complex index of deprivation and poverty comprising five dimensions: educational (EMPI), childhood/youth (YMPI), employment (WMPI), health (HMPI), and dwelling (DMPI) poverty conditions. These dimensions contain information on 15 variables (EMPI: illiteracy, low educational attainment; YMPI: school non-attendance, school lag, barriers to childhood care services, child labor; WMPI: informal employment, economic dependency; HMPI: lack of health insurance, barriers to health services; DMPI: lack of access to safe water, inadequate disposal of human feces, poor housing construction, and critical overcrowding), which contribute percentage differently in each department according to the adjusted MPI, which reflects information on the number of multidimensionally poor people (incidence of poverty) and the proportion of deprivations that this group dares.[11] As the calculation of the MPI changes from one year to the next, we only use data for one year, meaning this covariate will only capture the spatial variation in the case-fatality risk.[15] The childhood/young poverty dimension YMPI was considered as baseline to avoid collinearity, as HMPI+EMPI+YMPI+WMPI+DMPI = 100%, represents the percentages of contribution to each dimension to the adjusted MPI of each department.[11] Variables definition and a detailed explanation of the calculation of the adjusted MPI can be found in the supplementary material S1.

### 2.2. Case-fatality ratio calculation

Due to the heterogeneity of the population, we calculate a standardized case-fatality ratio (SCFR) employing an indirect standardization method for each Colombian department.[12] Thus, as shown in the Table S2 of supplementary material, we calculated age-specific expected deaths (E) for each age group in each department multiplying the values of the population projection for the year 2020 calculated using the 2018 population census,[13] by the case-fatality rate by age group previously established in Italy and China.[14] In this manner, we consider that areas with larger and more elderly populations are likely to exhibit a higher case-fatalities and avoid misleading comparisons using crude rates based on the population under study as a whole. Accordingly, the SCFR was computed for each department dividing the INS reported deaths (O) by the calculated expected deaths (E).

### 2.3. Bayesian Modeling

A conditional autoregressive (CAR) model and Markov chain Monte Carlo (MCMC) simulations were used to describe the variation in the SCFR, considering prior distributions for a set of spatial and temporal random effects.[15] This kind of model is usually proposed to analyze the epidemiological aspects of different disease dynamics.[15-20] Initially, a Poisson log-linear model was formulated as:

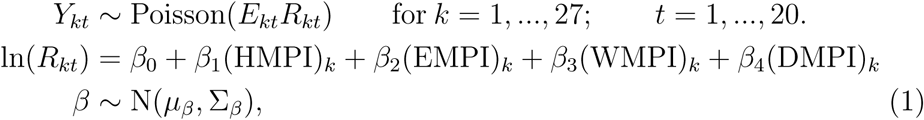

where *R_kt_* is the case-fatality risk relative to *E_kt_*, in the department *k* and week *t*. The number of deaths is denoted by **Y** = (**Y**_1_,…,**Y***_N_*)*_K_*_×_*_N_*, where **Y***_t_* = (*Y*_1_*_t_*,…, *Y_Kt_*) denotes the *k* × 1 column vector of observed deaths for all *K* departments for the week *t*. The vector of covariate regression parameters is denoted by *β*, and a multivariate Gaussian prior was assumed with mean *μ_β_* and diagonal variance matrix Σ*_β_*.

To check for the null hypothesis of spatial independence we used the Moran’s I statistic based on 10,000 random permutations of the data given by:

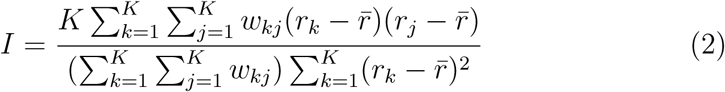

where 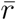 is the mean of each MPI, *r_k_* denotes MPI’s for the *k*th department and *r_j_* denote MPI’s at another department *j*. *W* = *(w_k_j*), is a *k* × *k* neighbourhood matrix, where *w_kj_* = 1 if departments (*k*, *j*) share a common border, and *w_kj_* = 0, otherwise.

Finally, we considered a spatio-temporal structure through a latent component for the department *k* and week *t* denoted by *ψ_kt_*, encompassing spatio-temporally autocorrelated random effects, and the complete set is denoted by *ψ* = (*ψ*_1_,…,*ψ_N_*), where *ψ_t_* = (*ψ*_1_*_t_*,…, *ψ_Kt_*). [19] Thus, adjusting equation 1,

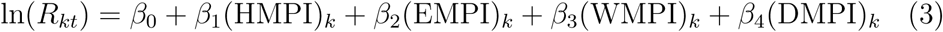

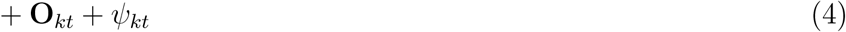

where a vector of known offsets is denoted by **O** = (**O**_1_,…, **O***_N_*)*_K_*_×_*_N_*, and similarly **O***_t_* = (*O*_1_*_t_*,..,*O_Kt_*) denotes the *k* × 1 column vector of offsets for week *t*. Then, the model was concluded by considering:

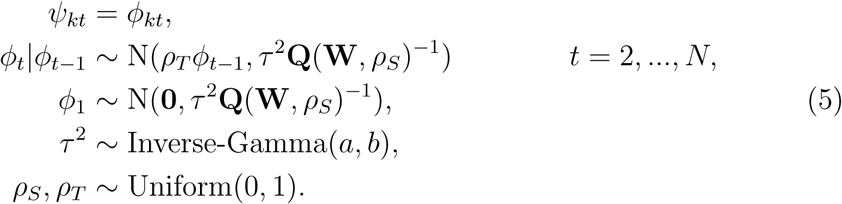

where, *ϕ_t_* — (*ϕ*_1_*_t_*,…, *ϕ_kt_*) is the vector of random effects for week *t* that evolve over time via a multivariate first order autoregressive process with a temporal autoregressive parameter *ρ_T_*. The temporal autocorrelation is thus induced via the mean *ρ_T_ϕ_t_*_-1_, while spatial autocorrelation is induced by the variance *τ*^2^**Q**(**W**, *ρ_S_*)^-1^. The precision matrix is given by **Q**(**W**, *ρ_S_*4)= *ρ_S_*(diag[**W**1] − **W**) + (1 − *ρ_S_*)**I**, where (**1**, **I**) is a *K* × 1 vector of ones and the *K* × *K* identity matrix respectively. Hence, the spatial autocorrelation is induced by the neighbourhood matrix **W**.

We generated MCMC samples from three independent Markov chains. Each chain was run for 1100,000 samples, of which 100,000 were removed as the burn-in period and the remaining 1000000 samples were thinned by 1000 to remove correlation amongst the samples. This leaves 1000 samples for inference from each chain. All analyzes were conducted in R version Version 1.3.959,[22] using the ‘CARBayesST’ package version 3.1,[23] following the recently published manual on spatio-temporal disease risk modeling in R using Markov chain Monte Carlo simulation and the CARBayesST package.[15]

The effects of each MPI on the COVID-19 SCFR were quantified as relative risks, for a fixed increase ξ in each covariates value. Thus, the relative risk for a ξ increase in each HMPI is computed by:

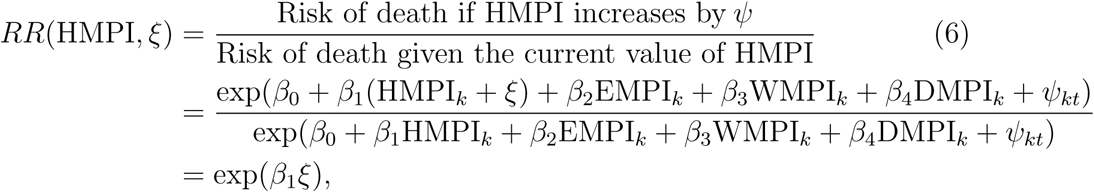

and equivalently calculated for EMPI, WMPI, DMPI and YMPI.

## 3. Results and Discussion

### 3.1. Standardized case-fatality rate

Figure 1A shows the SCFR for the 27 departments from Colombia with deaths reported from week 12 (from March 16) to week 31 (to August 02) of 2020. An SCFR equals 1, which means than the observed fatality risk is equal to the expected one, was not observed in any department. La Guajira, Putumayo, Norte de Santander, Córdoba, and Cesar, represent the highest case-fatality risk departments for COVID-19. La Guajira, in northern Colombia, exhibited the highest case-fatality risk with a mean SCFR of 16.2, which corresponds to an increasing of 1620% in the observed COVID-19 deaths compared to the expected value over the 20 weeks study period. Casanare, in eastern, and Antioquia in northwestern Colombia were the departments with the lowest case-fatality risk with a mean SCFR of 1.1 and 1.5, which corresponds to a 10% and a 50% higher risk, respectively.

For each department, the MPI is divided into the percentual contributions of each dimension (education, childhood/youth, employment, health, and dwelling), as described in Figure 1. These percentage contributions range from 5% to 40%, where the highest ones are typically given by the education and employment dimensions (together correspond to more than 50%), as shown in Figure 1C and Figure 1E, respectively. The relation between the variation of the percentage contributions of each dimension and the temporal average SCFR is not visually apparent. For instance, Antioquia (index 2 in Figure 1A) and Córdoba (index 13), which have one of the lowest (1.5) and highest (10.28) SCFR, respectively, are characterized by similar percentage contribution of the employment, health, and dwelling poverty dimensions. In contrast, in Santander (index 24) and Boyacá (index 6), which have the same SCFR (e.g., approximately 4), the percentage contributions of these poverty dimensions are also similar. Additionally, some spatial trends can be observed in the MPI dimensions. For instance, from western to eastern, Santander (index 24), Boyacá (index 6), Casanare (index 9), Meta (index 18), Cundinamarca (index 14), Bogotá DC (index 4), Caldas (index 7), Tolima (index 26), Cauca (index 10), and Nariño (index 19) have a percentage contribution of the health dimension to the MPI around 20% (Figure 1B). However, this trend is not followed by the SCFR (Figure 1A), suggesting that the health dimension by itself does not explain the SCFR heterogeneity and therefore case-fatalities inequality.

**Figure 1:**
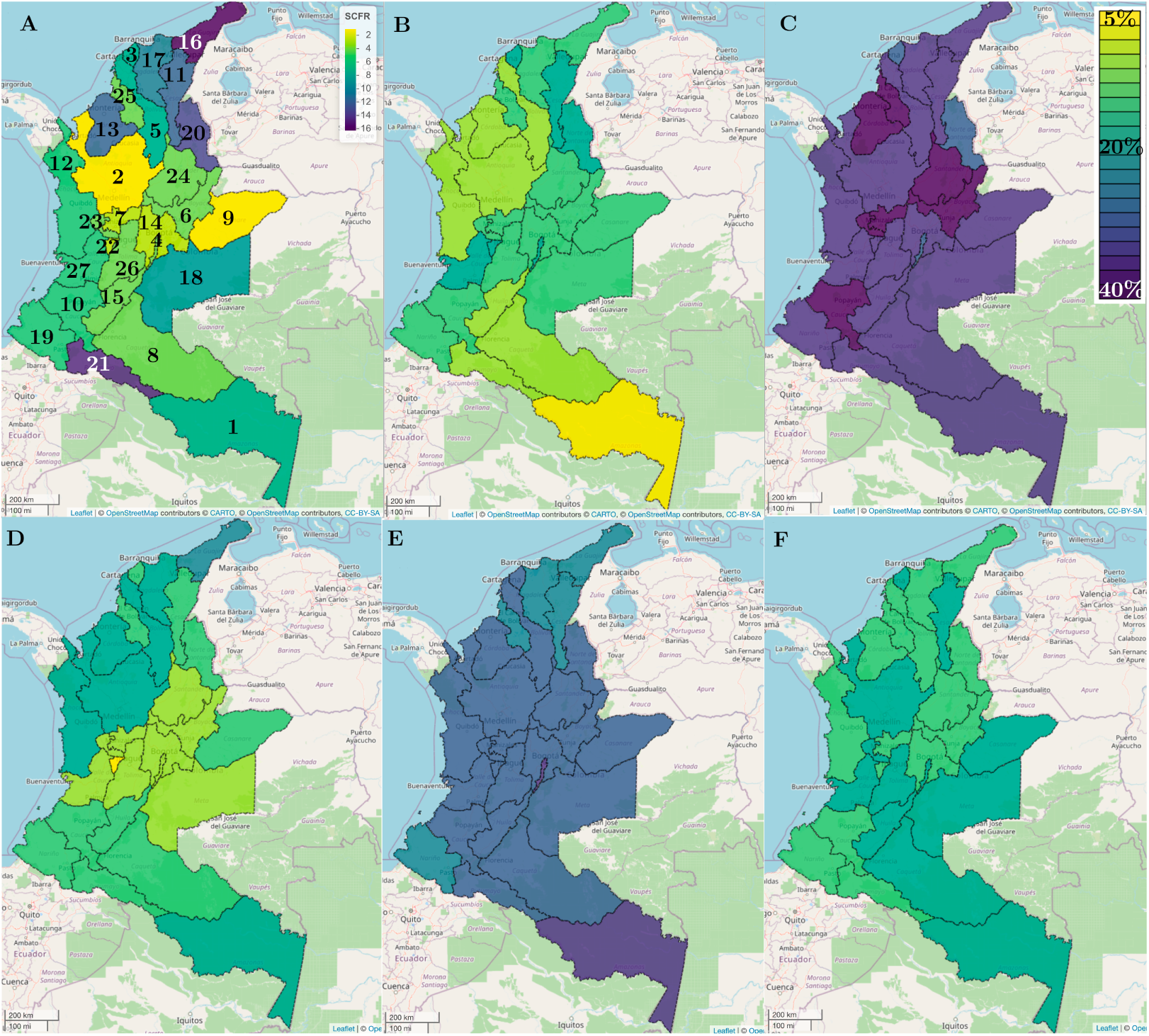
Spatial map of the A) average temporal SCFR in the study period, B) health, C) educational, D) dwelling, E) employment and F) childhood/youth poverty dimensions of the Multidimensional Poverty Index. Indexes correspond to 1:Amazonas, 2:Antioquia, 3:Atlántico; 4:Bogotá DC.; 5:Bolivar; 6:Boyacá; 7:Caldas; 8:Caquetá; 9:Casanare; 10:Cauca; 11:Cesar; 12:Chocó; 13:Córdoba; 14:Cundinamarca; 15:Huila; 16:La Guajira; 17:Magdalena; 18:Meta; 19:Nariño; 20:Norte de Santander; 21:Putumayo; 22:Quindio; 23:Risaralda; 24:Santander; 25:Sucre; 26:Tolima; 27:Valle del Cauca.

### 3.2. Bayesian multivariate Poisson regression

Before incorporating spatio-temporally autocorrelated random effects into the model, we modeled the data with a Bayesian multivariate Poisson loglinear model. Table 1 shows the results of this approximation. From the point estimates and 95% confidence intervals, in addition to the health dimension, notably all other dimensions exhibit significant effects on the COVID-19 case-fatality rate, with increases in the contribution of the health, employment, dwelling, and education dimensions to the MPI being associated with an increased risk of death for COVID-19 in Colombia.

**Table 1:** Intercept and coefficients in the Poisson log-linear model.

|  | Estimate | 2.5% | 97.5% | Std.Error | z.value | Pr(> \|z\|) |
| --- | --- | --- | --- | --- | --- | --- |
| (Intercept) | -8.885 | -10.553 | -7.217 | 0.851 | -10.443 | <2e-16 |
| HMPI | 0.208 | 0.185 | 0.231 | 0.012 | 17.773 | <2e-16 |
| WMPI | 0.084 | 0.062 | 0.106 | 0.011 | 7.502 | <6.3e-14 |
| DMPI | 0.167 | 0.147 | 0.187 | 0.010 | 16.460 | <2e-16 |
| EMPI | 0.092 | 0.073 | 0.111 | 0.009 | 9.668 | <2e-16 |

AIC: 9046.7

Since the sum of the contributions of the dimensions to the MPI is equal to 100%, the interpretation of Table 1 must take into account the childhood/youth dimension as baseline. Thus, for each 10% higher health dimension contribution (and hence 10% lower childhood/youth dimension contribution) and all else dimensions invariable, risk of death by COVID-19 is higher by 2.08 (95%CI 1.85-2.31). Similarly, for each 10% higher dwelling dimension contribution COVID-19 death risk is higher by 1.67 (95%CI 1.47-1.87). Additionally, for each 10% higher employment or education dimensions contribution COVID-19 death risk is higher by approximately 1. This shows that factors associated with the dwelling dimension, eg. lack of access to safe water, inadequate disposal of human feces, poor housing construction, and overcrowding, have a contribution to the SCFR similar to the one produced by the health dimension, made up of access to health services and lack of health coverage. Our analysis reveals that even without considering spatial autocorrelation, the poverty dimensions contributing to MPI allow us to predict the COVID-19 case-fatality risk.

### 3.3. Bayesian spatio-temporal modeling

The Moran’s test for the presence of spatial autocorrelation in residuals from the previous estimated non-spatio-temporal model, based on 10,000 random permutations of the data, suggests evidence of spatial autocorrelation from weeks 14 to 31 (fourth to twenty-first consecutive week with reported COVID-19 cases in Colombia) after accounting for the covariate effects, with a Moran’s I value between 0.13 and 0.46 and a p-value less than 0.05. The temporal trend of the SCFR for each department can be visualized in Figure S3 of the supplementary material. Thus, to quantify the evolution of the spatial pattern in the case-fatality rate over time, we fitted a spatially autocorrelated first-order autoregressive Bayesian model.

Table 2 provides a model description of the output and the model fitting criteria deviance information criterion (DIC). In addition to the median and 95% confidence intervals, Table 2 contains the effective number of independent samples (n.effective), and the result of Geweke diagnostic,[26] an MCMC convergence diagnostic that should lie between -2.0 and 2.0 to indicate convergence. The output shows that considering the childhood/youth dimension as baseline, all covariates i.e health, dwelling, employment, and education dimensions, exhibit positive relationships with case-fatality risk. Furthermore, the spatial *ρ_S_* and temporal *ρ_T_* dependence parameters exhibit that both spatial and temporal autocorrelation is present in these data after adjusting for the covariate effects.

**Table 2:** Posterior quantities for selected parameters and DIC of the autoregressive CAR model

|  | Median | 2.5% | 97.5% | n.effective | Geweke.diag |
| --- | --- | --- | --- | --- | --- |
| (Intercept) | -9.158 | -13.093 | -1.782 | 42.6 | 0.5 |
| HMPI | 0.188 | 0.115 | 0.226 | 27.4 | -0.5 |
| WMPI | 0.102 | 0.011 | 0.1681 | 53.6 | -0.1 |
| DMPI | 0.162 | 0.109 | 0.205 | 22.9 | 1.6 |
| EMPI | 0.092 | 0.039 | 0.162 | 135.2 | 0.1 |
| $\tau^2$ | 0.124 | 0.092 | 0.173 | 717.9 | 0.4 |
| $\rho_S$ | 0.849 | 0.662 | 0.931 | 924.9 | -1.3 |
| $\rho_T$ | 0.941 | 0.891 | 0.975 | 483.1 | -1.1 |

DIC= 2342.0

Figure 2 shows the average temporal case-fatality risk across the departments of Colombia for each week and MCMC sample, yielding the posterior distribution of these spatial averages for each week. The figure shows a clear trend in case-fatality risk, with a maximum between weeks 12 and 19 and a decrease from week 27. Although a temporal decrease is evident, the relative risk of dying by COVID-19 in Colombia is still 200% higher than the established case-fatality risk based on the COVID-19 dynamics in Italy and chain.

**Figure 2:**
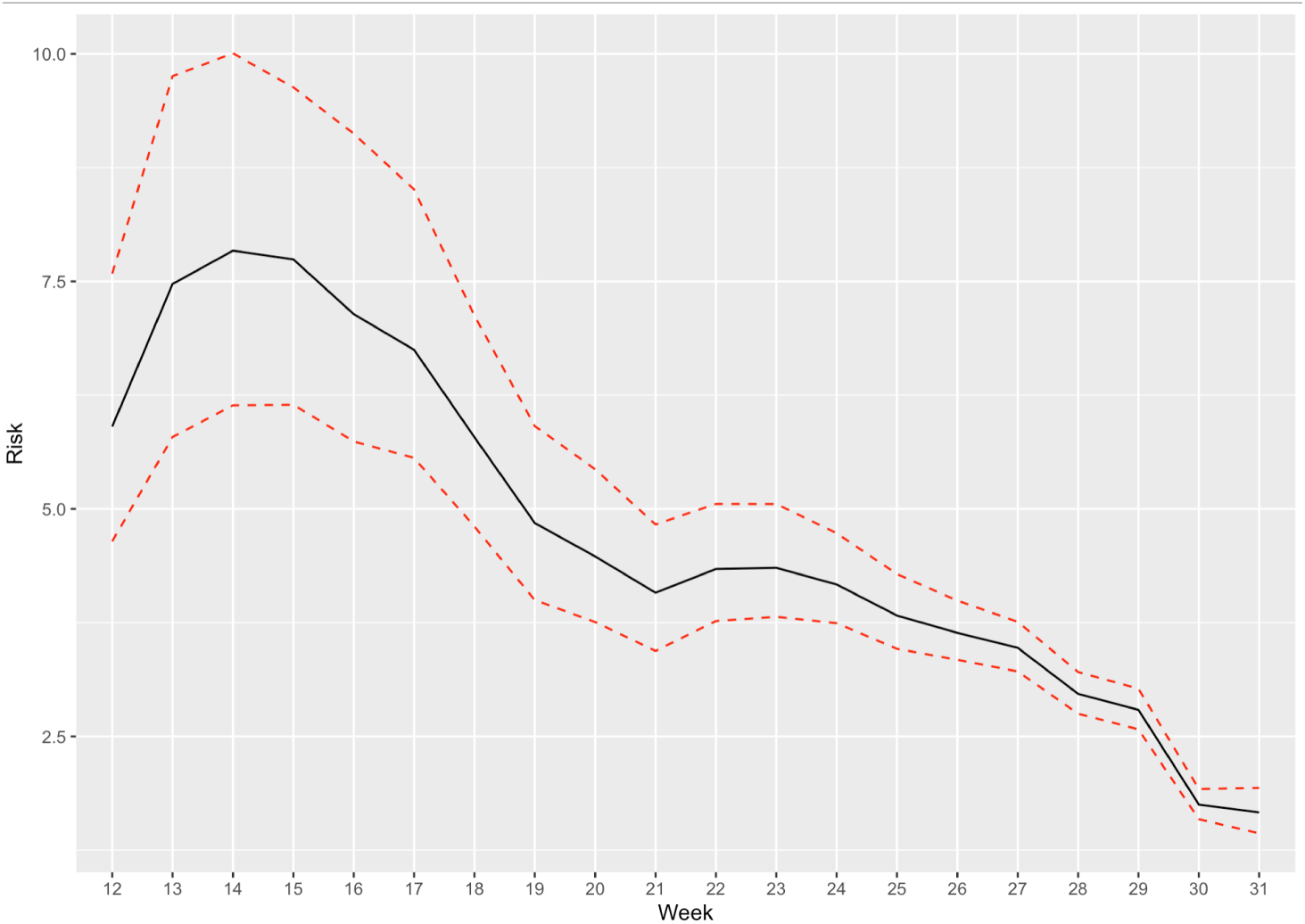
Posterior median and 95% credible interval for the temporal trend in case-fatality rate of COVID-19 in Colombia.

Estimated relative risks for the regression parameters show that the posterior relative risk for the HMPI dimension was 1.89 (95%CI=1.43-2.48), thus, in departments where the health dimension contributes in a greater proportion to the adjusted MPI the COVID-19 case-fatality risk increase by 189%. DMPI factors were also significantly related to COVID-19 case-fatality risk, with a posterior relative risk of 2.01 (95%CI=1.37-2.63), which suggests that in departments where the dwelling dimension contributes in a greater proportion to the adjusted MPI through the lack of access to safe water, inadequate disposal of human feces, poor housing construction, and critical overcrowding, the case-fatality risk for COVID-19 is 201% higher than in departments without these factors. Furthermore, in regions with a higher contribution of employment dimension (informal employment and economic dependency) to the MPI, the COVID-19 case-fatality rate is higher by 123% (95%CI=1.02-1.47). Finally, educational factors are also associated with COVID-19 death risk, thus, departments with a higher contribution of school non-attendance, school lag, barriers to childhood care services, and child labor factors to the adjusted MPI are at increased risk of dying from COVID-19 with a relative risk of 1.21 (95%CI=1.03-1.49).

Figure 3 shows the spatial pattern in case-fatality risk using the posterior median risk from week 14 onwards. Since week 14, COVID-19 case-fatality risk varies throughout the weeks in the different departments, initially mainly affecting the Amazonas, the Pacific, and Caribbean coastal regions, and later affecting the central area of the country. It is important to note that throughout the study period there were few departments and weeks in which the risk of death was close to 1.

**Figure 3:**
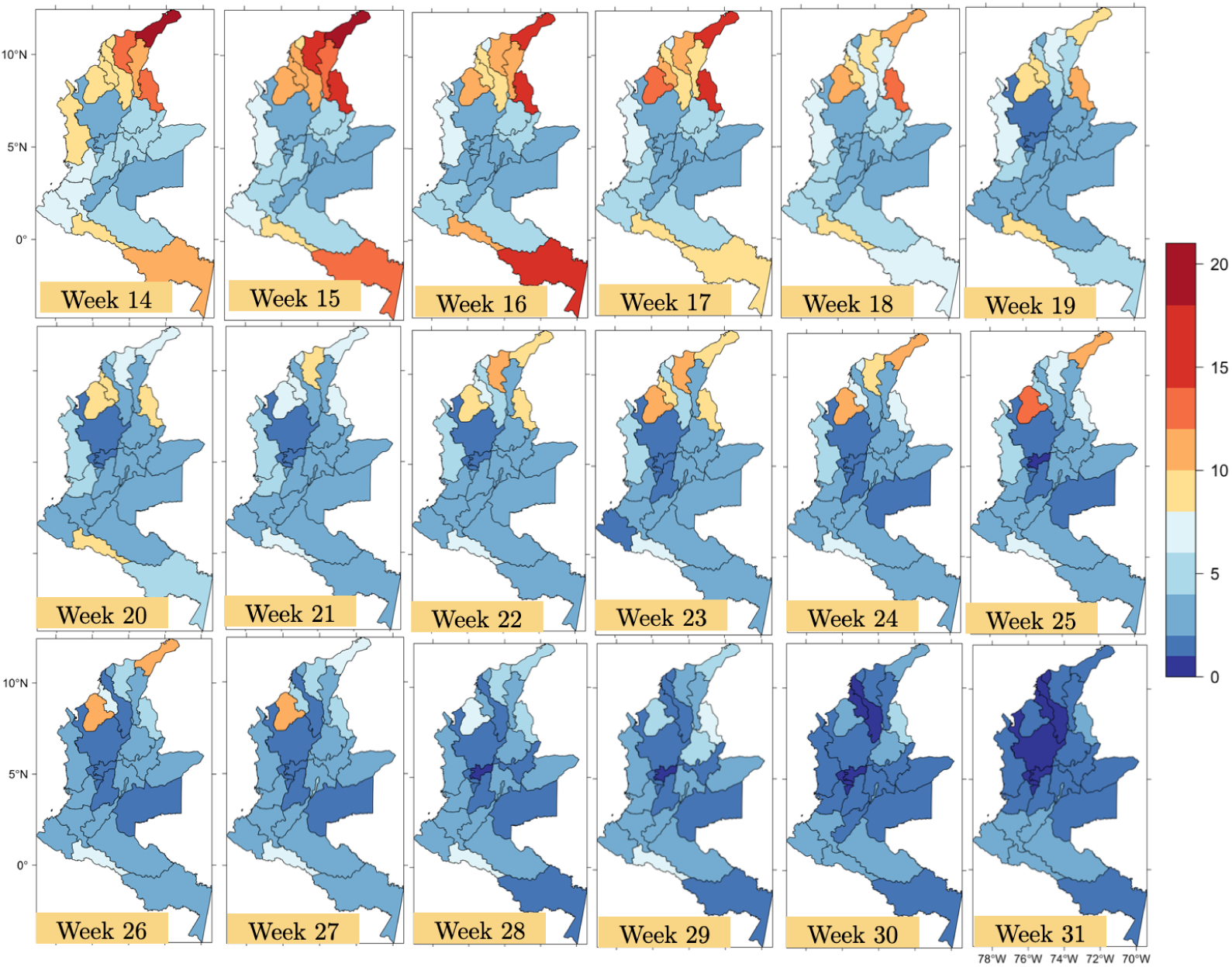
A) Spatial evolution of the posterior median estimated case-fatality risk from week 14 onwards.

COVID-19 case-fatality heterogeneity could be related to the level of self-reporting, since social groups with low socioeconomic indicators usually have lower self-reporting morbidity rates.[24,25] HMPI considers the definition of the barriers to health services indicator as the proportion of individuals who do not access to a public health service due to an illness that does not require hospitalization in the last 30 days.[11] Therefore, as HMPI is directly linked to self-reporting, we strengthen the importance of public strategies promoting the prompt self-reporting of symptoms associated with COVID-19 as a fundamental measure for the prevention of fatal cases. Furthermore, we verified that although the average COVID-19 testing turnaround time is similar across the country, the time from symptoms onset and the official notification/diagnostic is longer in social groups with low educational indicators (*r* = .6, *p* < .005). Furthermore, people living in more socio-economically disadvantaged regions have higher rates of the known clinical risk factors that increase the case-fatality risk of COVID-19.[8,27]

The provision of safe water, sanitation, and waste management are essential for preventing and protecting human health during the COVID-19 outbreak. [28] Basic prevention measures for the human-to-human transmission of COVID-19, such as handwashing [29], depend directly on the practice of hygiene measures in communities, homes, schools, marketplaces, and healthcare facilities. [28] Additionally, these measures, especially those for wastewater treatment, are fundamental and urgent when it is contemplated that the fecal-oral transmission of SARS-CoV-2 is possible [30] and even more so than wastewaters from hospitals or quarantine centers dedicated to COVID-19 treatments may contain SARS-CoV-2 RNA particles [31]. This is in agreement with the Bayessian analysis that spatially relate the COVID-19 case-fatality risk with the percentage contribution of the dwelling dimension to the MPI.

We consider the official INS report even containing biases inherent of underreporting and the number of diagnostic tests performed. However, we contemplated the spatial isotropic underreporting rate [10], which implies that although the values found for case-fatality risk are not definite, the described spatio-temporal trend is expected to continue. We verify that a spatially homogeneous increment in the observed cases, generates an adjustment of the intercept, however maintaining the importance of each MPI dimension as shown in the results of this work.

## 4. Conclusions

We provide evidence that the multidimensional poverty index significantly affects the inequalities in the COVID-19 case-fatality risk. Although educational and employment factors act as risk factors, the relationship was most robust and most consistent for health and dwelling aspects, showing higher risk associated in departments with lack of health insurance, low self-reporting, lack of access to safe water, inadequate disposal of human feces, poor housing construction, and critical overcrowding. These findings assist policy-makers in the spatial and temporal planning of strategies focused on mitigating fatality rates in most vulnerable communities and preparing for future pandemics by progressively reduce the factors that generate health inequality.

## Data Availability

Data supporting the findings of this study are freely available within the article references and its supplementary material.

## 5. Acknowledgment

We thank the Colombian Ministry of Science, Technology and Innovation for the MinCienciaton call, and the Vice-Rectory of Research and Transfer of Universidade de La Salle. We especially thank Alejandro Martinez Alvarez for his administrative support.

## 6. Funding

This work was funded by the Colombian Ministry of Science, Technology and Innovation - MinCiencias (MinCienciatón Project: 77464; Contract No. 362-2020. Project: “Modelización de intervenciones de Salud Pública del brote de COVID-19 en Colombia: efectividad e impacto epidemiológico y socio-económico de la toma de decisiones y medidas de mitigación”). The views expressed are those of the authors and not necessarily those of the MinCiencias.

## 7. Contributors

Conception and design: GP, CMA, and DST. Data acquisition: GP, CM, and DST. Statistical analysis and interpretation of the data: GP and CMA. Drafting of the manuscript: GP, CMA, DST, CMC, MP, JMM, JNB, SD, NP, JFPV, LVM, MR, LCV and EB. Critical revision for important intellectual content, discussion, review and final approval of the manuscript: all authors. Responsible for the overall content as the guarantors: GP, CM, DST and EB.

## 8. Competing interests

Authors declare no interests.

## 9. Data availability statement

Data supporting the findings of this study are freely available within the article references and its supplementary materials.

## Notes

### Competing Interest Statement

The authors have declared no competing interest.

